# Wastewater monitoring using a novel, cost-effective PCR-based method that rapidly captures the transition patterns of SARS-CoV-2 variant prevalence (from Delta to Omicron) in the absence of conventional surveillance evidence

**DOI:** 10.1101/2022.01.28.21268186

**Authors:** Taxiarchis Chassalevris, Serafeim C. Chaintoutis, Michalis Koureas, Maria Petala, Evangelia Moutou, Christina Beta, Maria Kyritsi, Christos Hadjichristodoulou, Margaritis Kostoglou, Thodoris Karapantsios, Agis Papadopoulos, Nikolaos Papaioannou, Chrysostomos I. Dovas

## Abstract

**Background:** The magnitude of Omicron SARS-CoV-2 variant of concern (VOC) global spread necessitates reinforcement of surveillance implementation. Conventional VOC surveillance based on genotyping of clinical samples is characterized by certain challenges related to available sequencing capacity, population sampling methodologies, and demands in terms of time, labor, and resources. Wastewater-based SARS-CoV-2 VOC surveillance constitutes a valuable supplementary practice, since it does not require extensive sampling, and provides information on the prevalence of the disease in a timely and cost-effective manner.

**Methods:** A highly sensitive real-time RT-PCR assay was developed, for targeted Omicron VOC detection and quantification in wastewater samples. The assay exclusively amplifies sequences with the S:Δ69/70 deletion, thus its performance is not hampered by the presence of the Delta VOC. The method was incorporated in the analysis of composite daily samples taken from the main Wastewater Treatment Plant (WWTP) of Thessaloniki, Greece from 1 December 2021 to 9 January 2022.

**Results:** The Omicron VOC was detected for the first time in samples from the Thessaloniki WWTP on 19 December 2021. In the following 10-day period, a rapid increase in Omicron sewage viral load was observed with an estimated early doubling time of 1.86 days. The proportion of the total SARS-CoV-2 load attributed to Omicron reached 91.09% on 7 January, revealing a fast Delta-to-Omicron VOC transition pattern. The detection of Omicron in wastewater preceded the outburst of reported (presumable) Omicron cases in the city by approximately 7 days.

**Conclusions:** The proposed wastewater-based surveillance approach enables rapid, real-time data acquisition on the Omicron VOC prevalence and transmission dynamics. Timely provision of these results to State authorities can readily influence the decision-making process for targeted public health interventions, including control measures, awareness, and preparedness.

## 1. Introduction

The widespread COVID-19 pandemic caused by the severe acute respiratory syndrome coronavirus 2 (SARS-CoV-2) is accompanied by the emergence and continuous reporting of several virus variants on a global scale [1]. Among them, Variants of Concern (VOCs) are of particular importance, as they are characterized by increased transmissibility, increased virulence/clinical disease presentation, or may hamper the performance of the available diagnostic assays, vaccines, therapeutics and public health measures [1,2]. Currently, five SARS-CoV-2 variants are designated as VOCs. The Alpha VOC (Pango lineage B.1.1.7) became the predominant SARS-CoV-2 variant worldwide in early January 2021 [3]. Concomitantly, the Beta (lineage B.1.351) and Gamma (lineage P.1) variants were also identified and reported [4]. In May 2021 the Delta variant (lineage B1.617.2) subsequently became dominant, accounting by November 2021 for nearly all infections globally [5,6].

The Omicron variant (lineage B.1.1.529) was identified recently in multiple countries and was designated as a VOC in late November 2021 [2], as it possesses many mutations, some of which may be linked with immune escape potential and enhanced transmissibility [7]. The Omicron VOC is driving an unprecedented surge of infections globally and initial reported spread was among younger individuals who tend to have more mild disease [8]. In this regard, early warning and surveillance of Omicron emergence is challenging, especially in areas where the incidence of Delta VOC is high.

The global phylogeny of Omicron shows the presence of 3 subclades; BA.1, BA.2, and BA.3. All Omicron variant strains, except for those within subclade BA.2 have an in-frame 6 nucleotide deletion at the Spike (S) protein gene, resulting in the absence of amino acids 69 and 70 (S:Δ69/70). As of 15 December 2021, BA.1 accounts for 99% of the sequences submitted to GISAID. Additionally, >95% of the Omicron variant sequences reported include the S:Δ69/70 [7]. Amino acids 69 and 70 are located in the N-terminal domain of the S1 fragment and it has been hypothesized that deletion of these residues may allosterically change S1 conformation [9]. This genomic trait also affects the performance of COVID-19 diagnostic tests that are based on the S-gene target failure (SGTF), as it has been previously described for the Alpha VOC, which also possesses the deletion.

Whole genome sequencing (WGS) via the use of the next-generation sequencing (NGS) technology constitutes the reference methodology for the detection of SARS-CoV-2 variants. However, the turn-around time of this approach is long and thus, its capability to help assist towards rapid public health responses is limited [10]. Besides being time-consuming, the universal application of NGS is also currently not feasible, since it is characterized by high running costs, requires highly trained personnel and is labor-intensive. Lastly, its capacity is limited, especially in samples with low viral loads, where failures are also frequent [11]. Consequently, alternative methodologies for the rapid detection of mutations and VOCs have been proposed by the ECDC, including the use of RT-PCR-based assays [10]. So far, such real-time RT-PCR-based methodologies were developed by our team to facilitate identification of critical SNPs associated with Alpha, Beta/Gamma and Delta VOCs, and the performance of the assays was subsequently evaluated in previously characterized human and veterinary clinical specimens [12,13].

Although variant identification in clinical samples is of importance, not all individual samples are expected to undergo variant typing in a resource-preservation effort, and thus, the analyzed samples may not represent the target population. In parallel with conventional surveillance, wastewater surveillance is being applied extensively for the monitoring of SARS-CoV-2 loads and mutations. It has been shown that SARS-CoV-2 RNA concentrations in sewage correlate with COVID-19 incidence in the respective areas, and for this reason, tracking VOCs through wastewater testing comprises an appealing and arguably a more accurate approach [14]. The common approach to monitor SARS-CoV-2 in wastewater is through the use of real-time RT-PCR, accompanied by NGS to identify and quantify mutations and lineages [15,16]. However, the use of sequencing-based variant identification can be affected by the nature of sewage samples, which is likely to comprise a “pool” of genetic material from multiple variants shed from different individuals, that reduces the breadth of sequence coverage [17]. On the contrary, via the utilization of RT-PCR-based approaches, target mutations in wastewater can be identified very fast, even if the respective variants are present at very low concentrations [18]. As an example, the development and use of a droplet-digital RT-PCR assay has been described to monitor abundance of mutations present in the Alpha and Delta VOCs in wastewater settled solids [14].

Already from the beginning of the pandemic, the wastewater-based epidemiology group of the Aristotle University of Thessaloniki in Greece has been engaged in analyzing the viral load in daily wastewater samples taken from the Wastewater Treatment Plant of Thessaloniki [19]. The group has published a number of papers that go beyond the conventional monitoring of viral load and contribute to the reliability of determinations. Hence, a methodology has been introduced which rationalizes virus shedding rate data in wastewater with regards physicochemical parameters of wastewater to cope with virus loss by adsorption to sewage suspended particles [20]. The presence of chemical and biological species in wastewater that compete with virus particles in occupying adsorption sites on suspended solids along with topological complications of large scale sewage networks have been considered in a separate publication to assess the spatial distribution of virus shedding rate along the wastewater piping system of a city [21]. In addition, contrary to earlier simplistic statistical approaches, a rigorous model has been proposed to estimate the number of infected people from the viral load in wastewater based on detailed population balances of infected people dynamics and shedding rate variability during disease days [15].

Aim of the present study was the development of a novel, sensitive RT-PCR-based methodology, readily applicable in many diagnostic laboratories and capable of specifically detecting and quantifying the Omicron VOC in wastewater samples, even in the presence of high loads of the currently predominant Delta VOC. In late 2021, the city of Thessaloniki, Greece, was experiencing the 4^th^ COVID-19 epidemic wave that was attributed exclusively to the predominant Delta VOC. Reported cases peaked on 25 November and a gradual declining trend was observed since then. The first official evidence of Omicron VOC emergence in Thessaloniki indicated that until 20 December, Omicron had been identified in 3 clinical samples. This evidence became publicly available on 30 December 2021, by the National Genomic Surveillance Network for SARS-CoV-2 mutations [22]. By the end of 2021, the city experienced an unprecedented outburst of daily reported cases. Specifically, 4,920 cases were reported on 31 December 2021, which was the highest number of cases recorded since the beginning of the pandemic [23]. Thus, the credibility and usefulness of the developed methodology was assessed in a real-world scenario, as surveillance for the Omicron VOC was incorporated in the daily viral load monitoring in the wastewater of Thessaloniki.

## 2. Methods

### 2.1. Oligonucleotides

A primer pair was designed to amplify a 145 bp fragment of the SARS-CoV-2 S gene. The selected downstream primer (OmDo2: 5’-AGTAGTACCAAAAATCCAGCCTCT-3’, *Tm*: 63.6 °C) was designed to be specific to all 5 designated VOCs, including the Omicron. The upstream primer (OmUp: 5’-TCCAATGTTACTTGGTTCCATGTTATCTC-3’, *Tm*: 65.2 °C) targets the region that contains the 6-nucleotide deletion associated with S:Δ69/70 (nucleotide sequence TACATG; positions 21765-21770 on GenBank acc. No. NC_045512, isolate “Wuhan-Hu-1). The design of this primer was based on our previously developed real-time PCR assay for the specific detection of wild-type strains of lumpy skin disease virus based on a deletion that differentiates them from homologous vaccine strains [24]. More specifically, in our case, the OmUp primer can hybridize the respective target genomic sequences of Omicron VOC strains that possess the deletion (subclades BA.1 and BA.3). Since this deletion is also a feature of the Alpha VOC, as already mentioned, the combination of the OmDo2 and OmUp primers is expected to amplify Alpha-specific sequences as well, despite the T/C mismatch present between the upstream primer and Alpha sequences (Figure 1).

**Figure 1.**
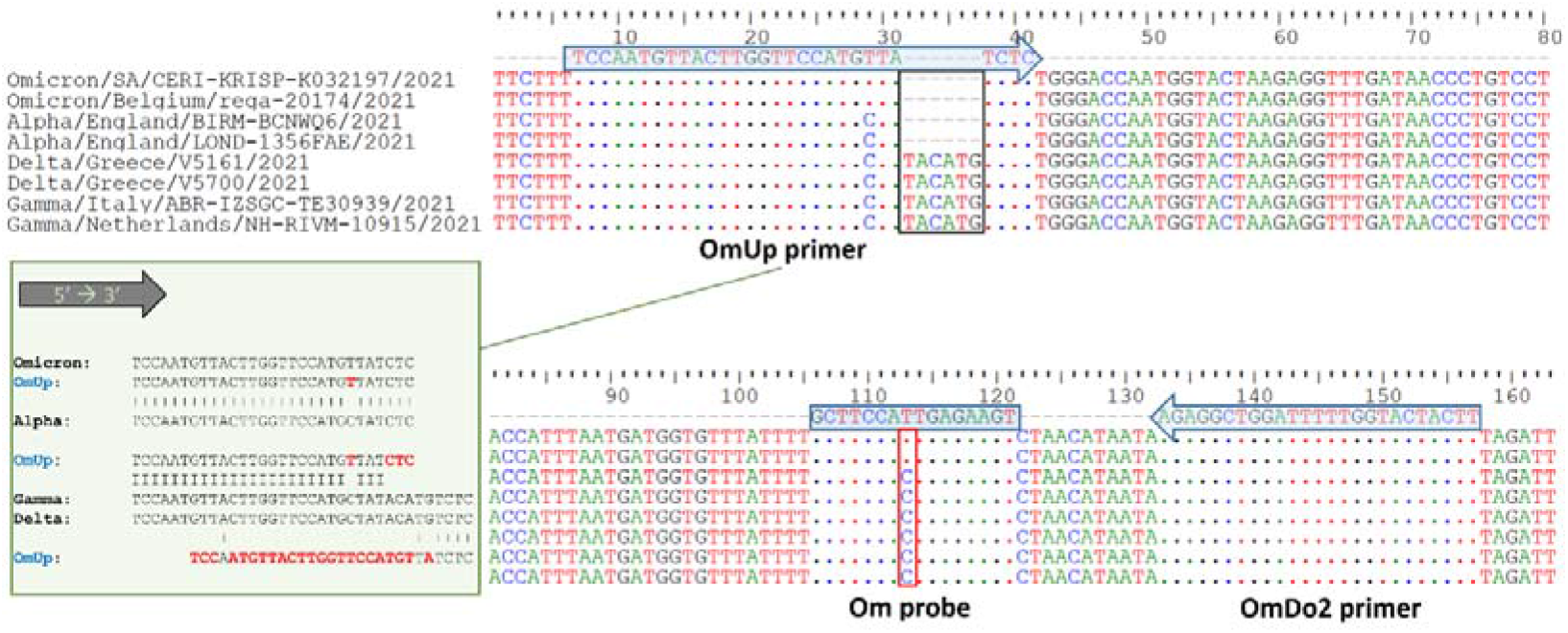
Multiple alignment of selected Omicron, Alpha, Delta and Gamma SARS-CoV-2 variant S gene sequences, indicating positions of primers (blue arrows) and TaqMan probe (blue rectangle). The 6-nucleotide deletion (dashes) targeted by the OmUp primer is indicated by a black rectangle. The C/T mismatch associated with the S:T95I amino-acid substitution in the probe-binding position is shown by a red rectangle. Inset: graphical display of the hybridization complexes between the upstream primer (OmUp) and SARS-CoV-2 VOCs (red: mismatches)

As Delta is the currently predominant variant worldwide, the assay’s design was focused to enable preferable amplification of Omicron VOC targets and minimize to the highest degree possible the amplification of Delta (along with Beta and Gamma that present similar characteristics at the respective upstream primer-hybridization regions). Additional primer sequence analysis was performed using the DINAMelt software [25] to estimate, at the annealing temperature of 58 °C, the mole fraction the OmUp primer hybridized to the target sequences of the designated VOCs [12,13,24].

A 6-carboxyfluorescein (FAM)-conjugated locked nucleic acid (LNA) TaqMan probe (OmProbe: 5’-FAM-ACTT+CTC+A+**A**+TGGAA+GC-IBFQ-3’, *Tm*: 62.5 °C) was designed to enable fluorescence detection. A mismatch position (bold) was included in this particular probe to provide the ability to discriminate variants based on the detection of the S:T95I amino-acid substitution (C/T nucleotide mutation) which is present in the Omicron VOC strains [4]. The effects of LNA modifications on mismatch discrimination were taken into consideration during probe design [26,27]. The length of the probe was also kept short (16 nucleotides) to improve mismatch discrimination. The philosophy behind the design of LNA TaqMan probes for SARS-CoV-2 VOC typing based on RT-PCR-based SNP identification has been presented in detail in our previous works [12,13]. Via this approach, discrimination between the Omicron and the Alpha is feasible on the bases of the obtained fluorescence.

### 2.2 Real-time RT-PCR assay

The reactions (20-μl) were comprised by: EnzyQuest’s One-step RT-qPCR kit (Product No.: RN010; EnzyQuest P.C., Heraklion, Greece), 4 mM Mg^2+^, the primers OmUp and OmDo2 at concentrations of 0.2 μM each; the OmProbe at concentration of 0.15 μ M, and 4 μl of sample RNA extract. Optimization of reactions took place on a CFX96 Touch Real-Time PCR Detection System (Bio-Rad Laboratories, Hercules, CA, USA). The following thermal cycling conditions were applied: 55 °C for 15 min (reverse transcription), 94 °C for 15 min (inactivation of reverse transcriptase and activation of Taq polymerase), and 48 cycles in 2 steps: i) 94 °C for 10 s (denaturation), and ii) 58 °C for 35 s (combined annealing/extension), followed by FAM fluorescence measurement. Fluorescence data acquisition and analysis were performed using the CFX Maestro Software (v4.1; Bio-Rad Laboratories, Hercules, CA, USA).

### 2.3 Analytical sensitivity, specificity, and diagnostic performance

For the evaluation of the analytical sensitivity parameters, a patient-derived SARS-CoV-2-positive RNA extract was used and the presence of the Omicron VOC in it was verified by NGS analysis. The RNA extract was subsequently subjected to real-time RT-PCR-based SARS-CoV-2 quantification, utilizing the N2 protocol proposed by the CDC for the diagnosis of COVID-19 in humans, with the use of the synthetic single-stranded RNA standard “EURM-019” (Joint Research Centre, European Commission) as a calibrator. Based on the quantification results, the sample RNA was serially 10-fold diluted and dilutions representing from 15 × 10^4^ down to 15 viral RNA copies/reaction were prepared. Each prepared dilution was tested in triplicate, to determine the amplification efficiency and linear range of the developed assay. The RNA extract was further diluted at 25, 20, 15, 10, 5 and 2.5 copies/reaction, in a background of Delta VOC-containing RNA extract from sewage. These dilutions were tested in 20 technical replicates each. The limit-of-detection (LOD) was subsequently determined with 95% probability of detection, by applying probit regression analysis using the MedCalc v20 (MedCalc Software, Mariakerke, Belgium).

Evaluation of the assay’s specificity also involved testing of the currently predominant Delta VOC in sewage RNA extracts. This involved testing diluted RNA extracts from clinical samples containing 5 × 10^3^ genomic Delta VOC copies/reaction in 5 replicates each. These concentrations were chosen to deflect highest viral RNA concentrations in concentrated sewage RNA extracts. Higher concentrations were also tested. In addition, the specificity in sewage samples was evaluated by testing sewage samples (N = 22) obtained from Thessaloniki city over a 20-day period, between 9 and 30 November 2021, spanning the peak of the epidemic wave in Thessaloniki due to the Delta VOC. The input SARS-CoV-2 RNA copy number ranged between 484 and 1,662 copies/assay. Titration of the aforementioned samples was performed during the routine monitoring of SARS-CoV-2 viral load in the city’s wastewater [15]. Specificity testing also involved testing 4 patient RNA samples containing the Alpha variant. Amplification curves and the relative fluorescence unit values at the plateau phase were compared to Delta VOC RNA samples also tested by the assay.

### 2.4 Omicron VOC surveillance results in wastewater samples from Thessaloniki city

After its evaluation, the Omicron VOC detection assay was incorporated in the routine monitoring of SARS-CoV-2 viral load in wastewater samples of Thessaloniki city, which is being performed daily. Routine analysis of sewage samples of a given day is performed in 3 replicates as described previously in detail [15]. Subsequently, quantitative measurements of SARS-CoV-2 RNA in these samples undergo rationalization with respect to the adsorption of virus onto suspended solids and the role of wastewater organic load on it by means of a physicochemical model [20]. The results of rationalization are expressed in the form of a relative shedding rate, that is, the ratio of the shedding rate at any day versus a reference shedding rate value. Without loss of generality, as reference value was selected the average shedding rate – a low value but above the LOQ – during the epidemiologically calm period in Thessaloniki at the first week of October 2020. Since 10 December 2021, RNA extracts (3 per day) that were subjected to routine total SARS-CoV-2 load determination, also underwent specific analysis for the Omicron VOC, by testing each extract in 10 replicates, i.e., 30 reactions/day. Samples obtained between 1-9 December were retrospectively analyzed for Omicron as well. In positive reactions, the Omicron-specific standard curve was used for quantification. Quantified Omicron RNA extracts underwent rationalization [20] and viral loads were expressed as relative shedding rates.

To estimate the doubling time of SARS-CoV-2 Omicron VOC concentration in sewage, we computed the exponential fit curve using daily relative shedding rate sewage measurements for Omicron for the first 10 days since its detection. Doubling time (D) was calculated as follows: D = ln(2)/σ, where σ was the slope of the curve.

### 2.5 Ethics statement

The human samples used as controls for the determination of the performance of the developed method were collected at the emergency ward of the General Hospital of Larissa from patients, as part of the routine diagnostic procedure. The use of clinical samples for the real-time RT-PCR method evaluation was approved by the scientific committee of the hospital, in accordance with national legal and ethical standards (Decision No. 102/19-03-2021).

## 3. Results

The *in-silico* prediction of the melting profile of the OmUp primer-Delta VOC hybridization complex indicated that in the reaction’s annealing conditions, non-specific hybridization between the upstream primer and Delta VOC targets is expected at a percentage of approx. 60%. However, amplification of Delta (and Beta/Gamma) VOC templates is unexpected, due to the presence of 4 nucleotide mismatches at the 3’-end of the hybridization region of the upstream primer. Alternatively, the upstream primers may be able to hybridize at their 3’-end, but other than that the hybridization is largely unstable to promote elongation during PCR amplification (Figure 1). Thus, through the design of the OmUp primer, amplification of Delta and Beta/Gamma targets is largely not favored.

The amplification efficiency of the assay was determined to be 100%, with a linear range of over 5 log_10_ Omicron VOC RNA copies (Figure 2). The corresponding standard curve was also characterized by a R^2^ of 0.998 and a y-intercept of 42.507. The LOD for the detection of Omicron VOC in RNA from wastewater samples was determined at 21 copies/reaction.

**Figure 2.**
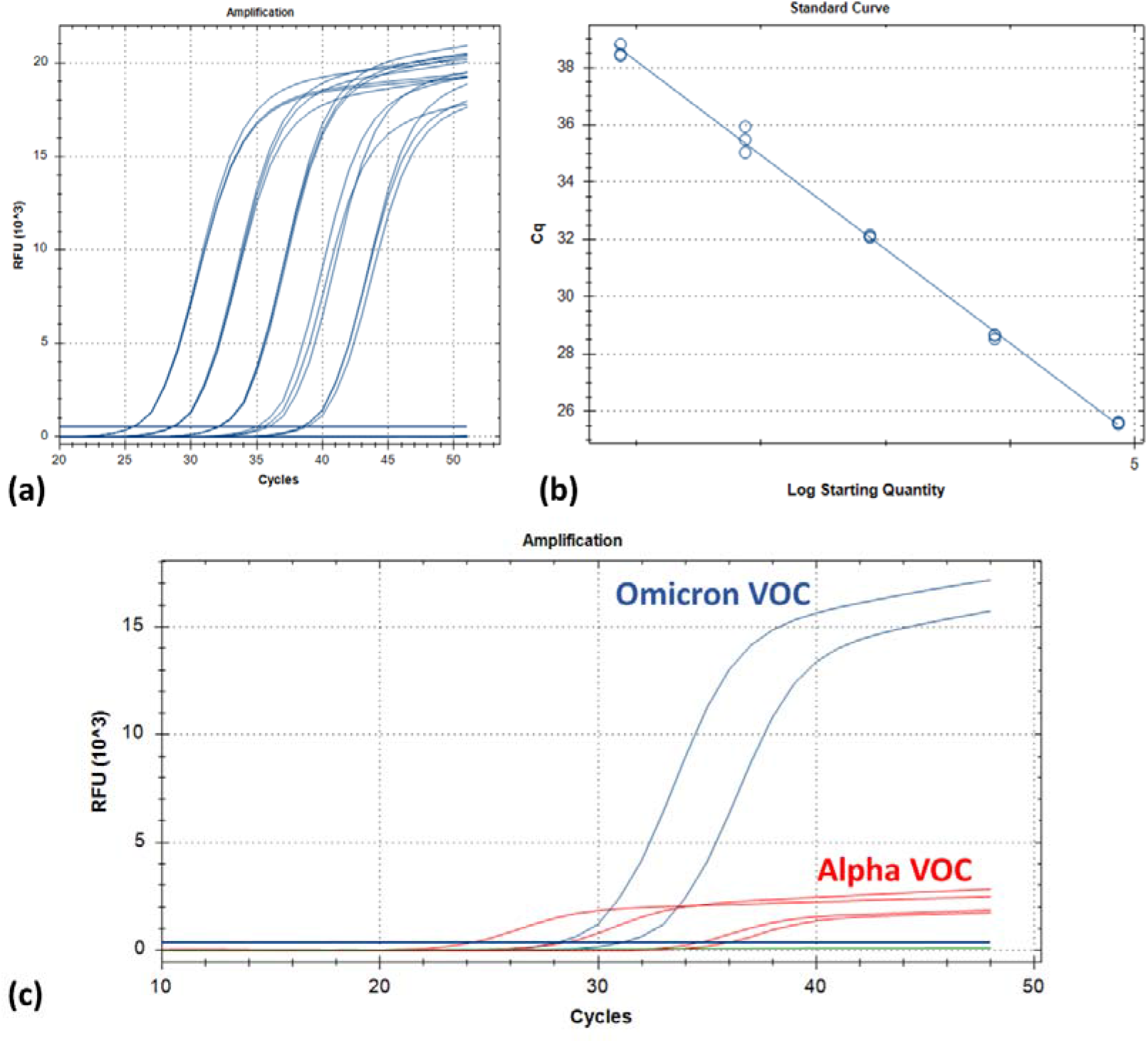
(a) FAM fluorescence amplification plots (blue) from testing 10-fold serial dilutions of the “Omicron” RNA standard, representing 15 × 10^4^ down to 15 viral RNA copies per reaction. Each dilution was tested in three replicates, to evaluate the amplification efficiency. The horizontal line indicates fluorescence baseline. RFU: relative fluorescence units; (b) the corresponding standard curve; (c) comparison of the fluorescence signals obtained by testing Alpha and Omicron VOC strains, which are both expected to be amplified by the upstream primer. The fluorescence signal for the Alpha VOC (red) is reduced compared to the respective signal for the Omicron VOC (blue) by over 80% RFU.

Results from testing four SARS-CoV-positive RNA extracts containing Alpha VOC strains revealed the detection of fluorescence. However, the respective amplification curves exhibited suppressed fluorescence signals compared to Omicron-specific curves (Figure 2). In detail, fluorescence signal for Alpha variant strains at plateau phase was considerably reduced by over 80% RFU (relative fluorescence units) compared to the respective signal for Omicron.

The *in-silico* prediction results about the absence of the Delta VOC detection were confirmed by the experimental evaluation of the specificity of the developed assay, where extracts with viral loads equivalent to 5 × 10^3^ genomic Delta VOC copies/reaction were tested, and fluorescence was not detectable. Delta VOC input RNA from clinical samples with very high high virus concentrations could be detected, although not efficiently, since a shift by approx. 13 cycles was observed in the Ct value obtained by the Omicron VOC assay, compared to the Ct value obtained when the same extract was tested with the CDC’s N2 assay (data not shown). Testing of sewage RNA extracts obtained from the wastewater treatment plant of Thessaloniki city between 9 and 30 November 2021 gave negative results, indicating that the highly abundant Delta VOC in sewage of that time was not detected.

After the incorporation of the assay in the sewage routine analysis of Thessaloniki wastewater (i.e. as of 10 December, along with the retrospective analysis of samples obtained between 1-9 December), the Omicron VOC was detected for the first time in the sample collected on 19 December 2021. Subsequently, a sharp increase in viral RNA concentrations was observed, as within a 10-day period, the levels of the Omicron VOC in Thessaloniki wastewater increased exponentially, from 6 RNA copies/μl (19 December) to 223 RNA copies/μl (29 December). Sewage viral load attributed to the Omicron VOC continued to increase at a lower rate, peaking at 675 RNA copies/μl (7 January 2022). The detection of the Omicron VOC in sewage coincided with a change in the overall trend of SARS-COV-2 RNA concentrations. Specifically, after the peak on 25 November, the SARS-CoV-2 load in sewage of Thessaloniki kept decreasing until December 18, i.e. a day before the detection of the Omicron VOC. The evolution of the viral load (expressed as relative shedding rates) corresponding to Omicron VOC over time is presented in Figure 3, along with the respective viral loads that correspond to the Delta VOC and the total measured SARS-CoV-2. The doubling time of the Omicron VOC load in sewage, based on measured relative shedding rate values, was estimated at 1.86 days, for a 10-day period (19-to-29 December).

**Figure 3.**
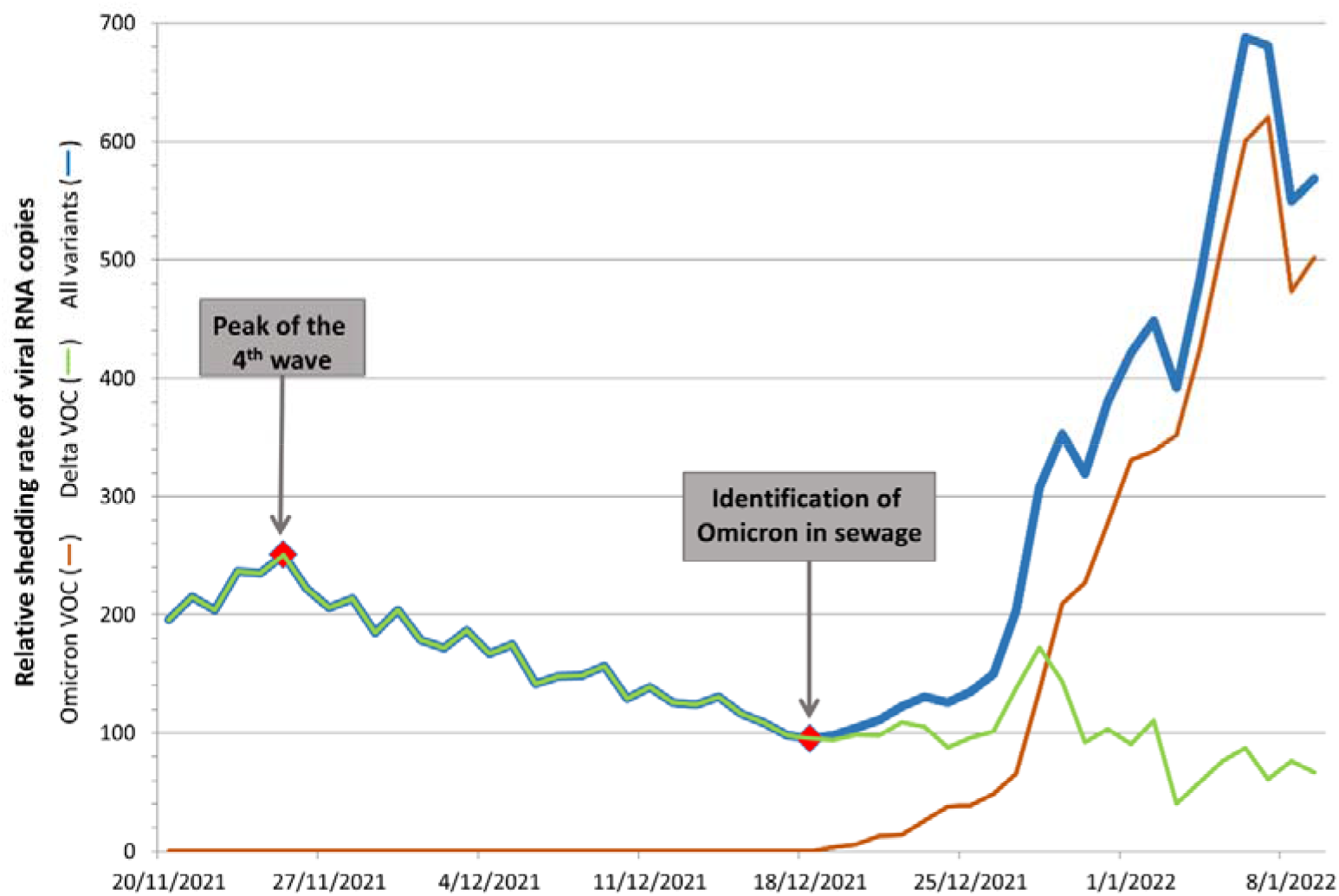
Fluctuations of wastewater SARS-CoV-2 viral load levels from samples obtained from the wastewater treatment plant of Thessaloniki. The relative shedding rates of total SARS-CoV-2, Omicron VOC and Delta VOC, are represented by the blue, brown and green line, respectively. The relative shedding rate of Delta VOC was calculated by subtracting Omicron VOC relative shedding rates from the total SARS-CoV-2 respective values.

The proportion of sewage RNA corresponding to the Omicron VOC also increased rapidly from 3.52% on 19 December 2021 to 89.78% on 3 January 2022, indicating a fast transition from the Delta VOC to Omicron VOC within the community of Thessaloniki city (Figure 4). The following period (4-9 January 2022) the above proportion remained stable with mild fluctuations indicating a degree of persistence of the Delta VOC at relatively low levels. The time series of SARS-CoV-2 load in wastewater against daily confirmed COVID-19 cases in the regional unit of Thessaloniki, as reported by the National Public Health Organization is presented in Figure 5. It is evident that, shortly after the identification of the Omicron VOC in sewage, an outburst of reported cases occurred. It can be also observed that the increases in relative viral shedding occur earlier than the increase in reported cases. While the exponential growth of cases becomes clear in the period between 27 and 31 December, the increasing trend of SARS-CoV-2 loads in sewage was observed as of 19 December, i.e. over a week earlier.

**Figure 4.**
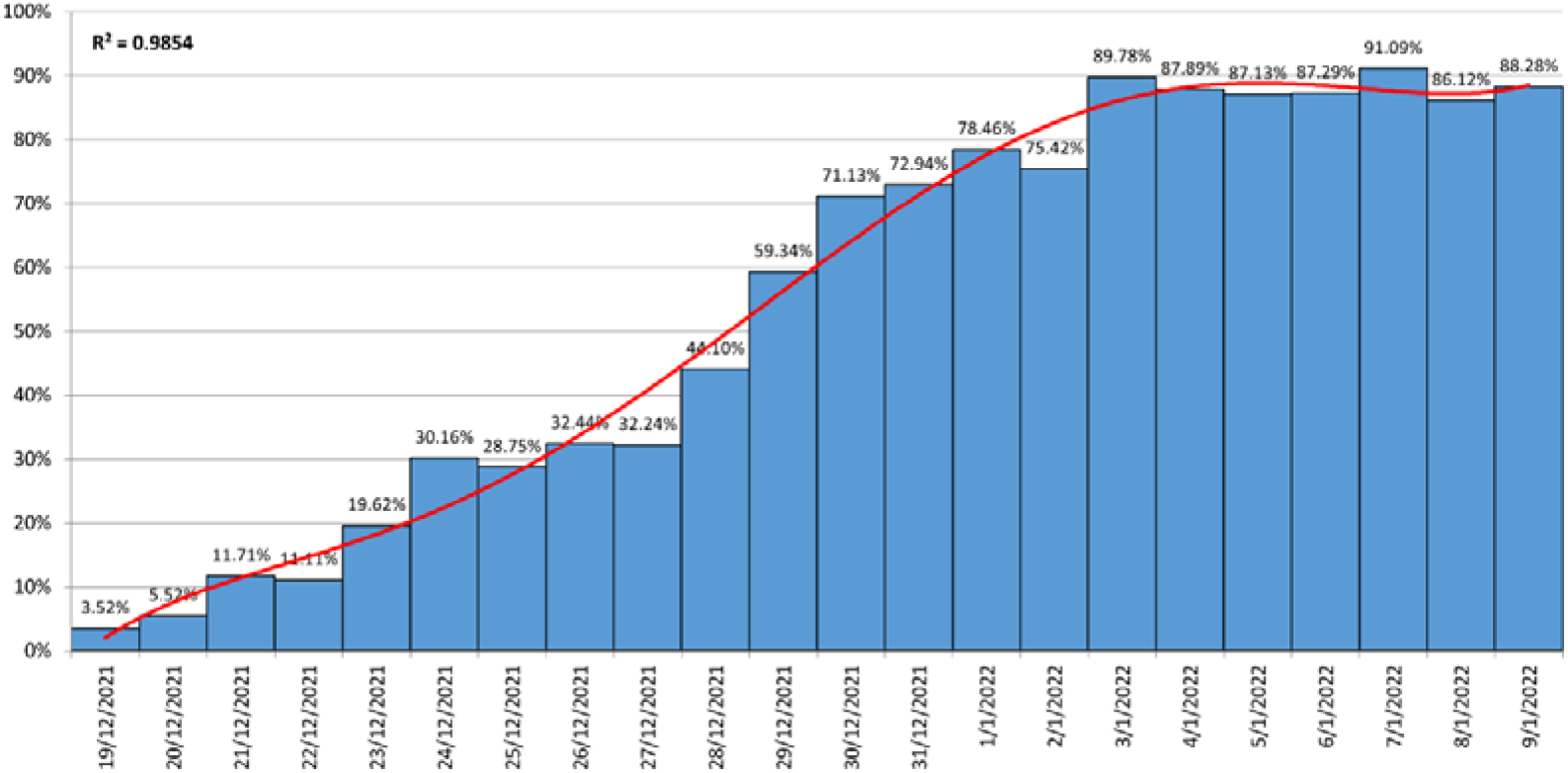
Proportion (%) of the total wastewater viral load corresponding to Omicron VOC from 19 December 2021 to 9 January 2022. Red line represents a polynomial fit curve

**Figure 5.**
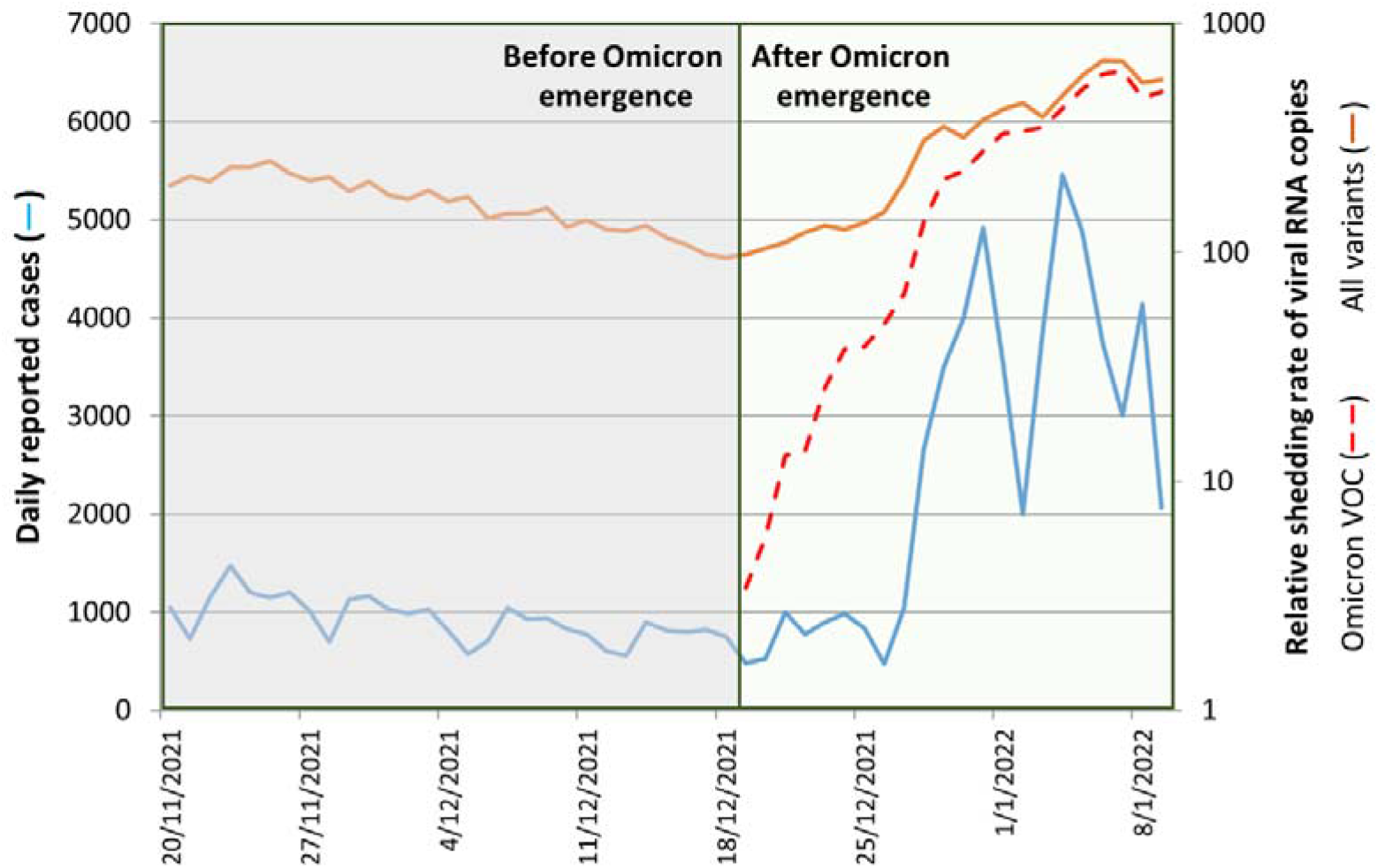
Wastewater SARS-CoV-2 levels and daily reported COVID-19 cases before and after the identification of Omicron in sewage from Thessaloniki city (20 November 2021 to 9 January 2022). Relative viral shedding (right axis) is expressed at a logarithmic scale. Daily reported cases (left axis; blue line) are presented in a linear scale.

## 4. Discussion

Although Delta is the predominant SARS-CoV-2 variant during the fourth COVID-19 wave in many countries worldwide, the novel Omicron variant was rapidly reported and designated as a VOC. This is since it harbors a high number of mutations in the S gene, and preliminary data from South Africa indicated that this variant is characterized by immune evasion capabilities, rapid transmissibility, increased disease severity and high re-infection rates [28,29]. The rapid spread in South Africa, particularly among younger age groups, placed WHO and global health systems on high alert [30]. It is also of concern that the Omicron VOC is rapidly spreading against a backdrop of ongoing Delta VOC transmission and high levels of natural immunity against the latter variant [31]. Since the rapid spread of the Omicron VOC at the first stages after its introduction in an area involves mostly young adults, the possibility to rapidly identify its emergence via the use of classical surveillance is consequently reduced.

The unprecedented magnitude of Omicron VOC expansion necessitates reinforcement in the implementation of surveillance utilizing molecular diagnostics techniques. Taking also into consideration the already discussed restrictions of the routine use of NGS for this purpose, an alternative approach was urgently needed to directly detect and quantify low levels of the Omicron VOC in sewage samples, rapidly and efficiently. Wastewater-based epidemiology comprises an alternative approach for monitoring the dynamics SARS-CoV-2 outbreaks with several benefits. Importantly, SARS-CoV-2 variant monitoring in sewage can allow a fine description of the variant spreading in the monitored population, as well as the dynamics of newly introduced variants [32]. Besides the analysis of sewage obtained from city treatment plants, wastewater analysis from incoming flights could also be utilized to monitor introductions of the variant [33]. So far, variant monitoring in wastewater is applicable mostly via the use of the NGS methodology. However, a relatively high quantity of input target RNA is required for this technique and considerable time is needed for sample analysis [16]. As introduction of rapidly expanding Omicron takes place in areas where the Delta is predominant, its early detection can be missed or delayed.

In this context, a highly sensitive real-time RT-PCR assay was developed for immediate and targeted detection and quantification of Omicron VOC strains in sewage samples. The assay exclusively amplifies sequences with the S:Δ69/70 deletion that is a trait of the majority of the Omicron VOC strains reported worldwide. Due to the fact that the deletion was exploited for the design of the upstream primer, amplification of the genome of variants that do not possess the deletion (e.g. of the Delta VOC) is not favored, thus addressing the issue of competition among amplicons corresponding to different VOCs. This ensures the applicability of the assay in use for the analysis of wastewater, as it comprises a composite sample containing genomes from various SARS-CoV-2 strains. Commercially available SGTF assays that are being used for diagnostics are different from the developed assay in this regard, as it has been shown that the Δ69/70 deletion is contained in the middle of the amplicon, meaning that all SARS-CoV-2 genomes are amplified, irrespective of the variant, and the SGTF relies on the inability of the probe to hybridize in deletion-containing amplicons [34]. Consequently, despite being useful for COVID-19 diagnosis by testing human specimens, those commercial assays are inappropriate for testing composite wastewater samples, as a novel variant can either be missed or incorrectly quantified when a different variant dominates. Evaluation of our assay in sewage samples indicated its robust performance in detecting low-copy numbers of the Omicron VOC, a feature that is of importance in order to detect the introduction of Omicron in a monitored area as early as possible. The presence of Delta VOC which was ubiquitous in the monitored wastewater samples in December 2021 did not affect the ability of the assay to detect and quantify Omicron. Additionally, fluorescence signals were not obtained even when much higher Delta VOC RNA copy numbers were tested, compared to those that were expected to be found in wastewater samples, e.g. at the concentrations detected in sewage samples from Thessaloniki during the peak observed in November 2021. Fluorescence corresponding to the Alpha VOC could be detected, as this variant also harbors the S:69/70 deletion. However, the fluorescence plateau values detected were considerably lower compared to those of the Omicron VOC. In addition, since the Alpha VOC has been completely displaced by the Delta VOC, the presence of the former variant in the monitored wastewater is unexpected.

To the authors’ knowledge, the present research reports for the first-time an early warning system based on wastewater-based surveillance that was able to specifically detect and rapidly estimate the viral load doubling time and frequency of Omicron VOC in a large Greek city, prior to its recognition through clinical testing, therefore rapidly enabling awareness and preparedness among the community. Upon the detection of the Omicron VOC in sewage of Thessaloniki on 19 December, the corresponding wastewater RNA levels were characterized by a sharp exponential growth. Omicron VOC influent levels showed a more than 60-fold increase within the first 10 days and over 100-fold increase within the first 15 days, with a short early doubling time. This observation is in agreement with estimates derived from surveillance data of reported cases, from countries where the Omicron VOC has prevailed. Early investigations from Gauteng Province of South Africa had estimated the early doubling time in the first 3 days after crossing the wave threshold of ten cases per 100,000 population at 1.2 days and at 1.5 days in the first week [31]. Since then, data on the Omicron transmission dynamics are rapidly accumulating, unanimously indicating significantly higher transmission rates compared to the Delta VOC. The World Health Organization has summarized recent evidence (until 17 December 2021) on the transmission of Omicron VOC suggesting a doubling time between 1.5–3 days, based on various studies and reports mainly from United Kingdom and South Africa [7]. Data from the UK Health Security Agency on 24 December 2021 also highlight the short doubling time that is < 2.5 days for various regions of England [35].

Our results indicate that the proposed method provided evidence on the increase of Omicron VOC prevalence significantly earlier than routine surveillance based on clinical samples. The detection of the Omicron VOC in wastewater influent of the wastewater treatment plant of Thessaloniki signified major changes in the total sewage SARS-CoV-2 viral load, immediately reversing the prevailing declining trends. This pattern change was later reflected in the epidemic curve. Reported SARS-CoV-2 cases in the regional unit of Thessaloniki started increasing exponentially approximately a week after the detection of Omicron in sewage, reaching unprecedented levels as of 31 December 2021. Thus, the present report provides a clear early warning example of wastewater-based epidemiology. It is interesting though, that this early warning potential was not clear in data before the emergence of the Omicron VOC. This can be attributed to the fact that Omicron is more prevalent in younger individuals who are more likely to escape diagnosis. Recent observations from the United Kingdom verify that the distribution of Omicron VOC by age currently differs markedly from the Delta VOC, as people aged 18-29 year-old have significantly higher infection rates with Omicron relative to Delta VOC [36].

We also observed a pattern of extremely fast Delta-to-Omicron transition in sewage SARS-CoV-2 load, as the proportion of Omicron VOC reached 60.2% within 10 days, and 91.09% within 19 days. The interpretation of clinical surveillance data reveals similar transition trends. According to UK Health Security Agency, the percentage of cases with SGTF in England increased from 2.47% (5 December 2021) to 67.3% (15 December 2021) in a 10 day period [37]. Moreover, the estimated proportions of sewage viral load attributed to the Omicron VOC are in line with results from the analysis of clinical samples from Thessaloniki, where the percentage of Omicron cases based on SGTF screening ranged from 67.5% to 85.0%, between 3 and 8 January 2022 (National Public Health Organization, personal communication). Thus, through the applied wastewater-based surveillance methodology, a similar pattern of Omicron VOC expansion was identified compared to surveillance data that are based on SARS-CoV-2 characterization in clinical samples. The proposed approach has a number of profound advantages over sequencing-based variant surveillance practices as it is significantly faster, less labor-intensive, inexpensive and more practical. The format of the assay enables its wide applicability in laboratories, with rapid turnaround of results, which comprises a critical characteristic of such assays that aim to influence the decision-making process for targeted public health interventions.

In conclusion, targeted Omicron VOC systematic surveillance in sewage samples accurately captured the patterns of Delta-to-Omicron transition within the examined community. The identification of the Omicron VOC in sewage may constitute a critical alarm point that can assist policy makers in regard to the timely application of response measures. The fact that community exposure to Omicron was identified approximately 7 days earlier than the outbreak of cases, together with estimates that the incubation period of the Omicron VOC is shorter [38], suggest that wastewater-based epidemiology can strengthen the preparedness against this VOC’s extremely rapid spread. The mitigation of the consequent epidemic waves is crucial, especially when the Delta VOC circulation is also prevalent, and together with outbreaks of seasonal influenza can lead to “winter’s twindemic” [39], applying overwhelming pressure to the healthcare systems. Finally, the proposed approach that targets deletions to selectively amplify VOCs can serve as a paradigm for monitoring the community spread of new variants that may emerge in the future.

## Data Availability

All data produced in the present work are contained in the manuscript

## References

[1] CDC, SARS-CoV-2 Variants of Concern, (2021). https://www.cdc.gov/coronavirus/2019-ncov/cases-updates/variant-surveillance/variant-info.html (accessed December 20, 2021).

[2] WHO, Tracking SARS-CoV-2 variants, (2021). https://www.who.int/en/activities/tracking-SARS-CoV-2-variants/ (accessed December 22, 2021).

[3] European Centre for Disease Prevention and Control (ECDC), SARS-CoV-2 variants of concern as of 22 December 2021, (2021). https://www.ecdc.europa.eu/en/covid-19/variants-concern (accessed December 24, 2021).

[4] CoVariants, CoVariants, (2021). https://covariants.org/shared-mutations (accessed December 18, 2021).

[5] H. Hwang, J.-S. Lim, S.-A. Song, C. Achangwa, W. Sim, G. Kim, S. Ryu, Transmission dynamics of the Delta variant of SARS-CoV-2 infections in South Korea, J. Infect. Dis. (2021). doi:10.1093/INFDIS/JIAB586.

[6] D. Tian, Y. Sun, J. Zhou, Q. Ye, The Global Epidemic of the SARS-CoV-2 Delta Variant, Key Spike Mutations and Immune Escape, Front. Immunol. 12 (2021). doi:10.3389/FIMMU.2021.751778.

[7] WHO, Enhancing Readiness for Omicron (B.1.1.529): Technical Brief and Priority Actions for Member States, 2021. https://www.who.int/docs/default-source/coronaviruse/20211217-global-technical-brief-and-priority-action-on-omicron_latest-2.pdf?sfvrsn=bdd8297c_9&download=true.

[8] WHO, Update on Omicron, (2021). https://www.who.int/news/item/28-11-2021-update-on-omicron (accessed January 17, 2022).

[9] S.S. Hwang, J. Lim, Z. Yu, P. Kong, E. Sefik, H. Xu, C.C.D. Harman, L.K. Kim, G.R. Lee, H.B. Li, R.A. Flavell, Cryo-EM structure of the 2019-nCoV spike in the prefusion conformation, Science. 367 (2020) 1255–1260. doi:10.1126/SCIENCE.ABB2507.

[10] European Centre for Disease Prevention and Control (ECDC), Methods for the detection and identification of SARS-CoV-2 variants, (2021). https://www.ecdc.europa.eu/en/publications-data/methods-detection-and-identification-sars-cov-2-variants (accessed August 18, 2021).

[11] B. Huang, A. Jennsion, D. Whiley, J. McMahon, G. Hewitson, R. Graham, A. De Jong, D. Warrilow, Illumina sequencing of clinical samples for virus detection in a public health laboratory, Sci. Reports 2019 91. 9 (2019) 1–8. doi:10.1038/s41598-019-41830-w.

[12] S.C. Chaintoutis, T. Chassalevris, S. Balaska, E. Mouchtaropoulou, G. Tsiolas, I. Vlatakis, A. Tychala, D. Koutsioulis, A. Argiriou, L. Skoura, C.I. Dovas, A Novel Real-Time RT-PCR-Based Methodology for the Preliminary Typing of SARS-CoV-2 Variants, Employing Non-Extendable LNA Oligonucleotides and Three Signature Mutations at the Spike Protein Receptor-Binding Domain, Life (Basel, Switzerland). 11 (2021). doi:10.3390/LIFE11101015.

[13] S.C. Chaintoutis, T. Chassalevris, G. Tsiolas, S. Balaska, I. Vlatakis, E. Mouchtaropoulou, V.I. Siarkou, A. Tychala, D. Koutsioulis, L. Skoura, A. Argiriou, C.I. Dovas, A one-step real-time RT-PCR assay for simultaneous typing of SARS-CoV-2 mutations associated with the E484K and N501Y spike protein amino-acid substitutions, J. Virol. Methods. 296 (2021). doi:10.1016/j.jviromet.2021.114242.

[14] A. Yu, M. Wolfe, T. Leon, D. Duong, L. Kennedy, S. Ravuri, K.R. Wigginton, A. Boehm, Estimating relative abundance of two SARS-CoV-2 variants through wastewater surveillance at two large metropolitan sites, (2021). doi:10.21203/RS.3.RS-1083575/V1.

[15] M. Petala, M. Kostoglou, T. Karapantsios, C.I. Dovas, T. Lytras, D. Paraskevis, E. Roilides, A. Koutsolioutsou-Benaki, G. Panagiotakopoulos, V. Sypsa, S. Metallidis, A. Papa, E. Stylianidis, A. Papadopoulos, S. Tsiodras, N. Papaioannou, Relating SARS-CoV-2 shedding rate in wastewater to daily positive tests data: A consistent model based approach, Sci. Total Environ. 807 (2022). doi:10.1016/J.SCITOTENV.2021.150838.

[16] N. Pechlivanis, M. Tsagiopoulou, M.C. Maniou, A. Togkousidis, E. Mouchtaropoulou, T. Chassalevris, S. Chaintoutis, C. Dovas, M. Petala, M. Kostoglou, T. Karapantsios, S. Laidou, E. Vlachonikola, A. Chatzidimitriou, A. Papadopoulos, N. Papaioannou, A. Argiriou, F. Psomopoulos, Detecting SARS-CoV-2 lineages and mutational load in municipal wastewater; a use-case in the metropolitan area of Thessaloniki, Greece, MedRxiv. (2021) 2021.03.17.21252673. doi:10.1101/2021.03.17.21252673.

[17] S. Wurtzer, P. Waldman, A. Ferrier-Rembert, G. Frenois-Veyrat, J.M. Mouchel, M. Boni, Y. Maday, V. Marechal, L. Moulin, Several forms of SARS-CoV-2 RNA can be detected in wastewaters: Implication for wastewater-based epidemiology and risk assessment, Water Res. 198 (2021). doi:10.1016/J.WATRES.2021.117183.

[18] W.L. Lee, M. Imakaev, F. Armas, K.A. McElroy, X. Gu, C. Duvallet, F. Chandra, H. Chen, M. Leifels, S. Mendola, R. Floyd-O’sullivan, M.M. Powell, S.T. Wilson, K.L.J. Berge, C.Y.J. Lim, F. Wu, A. Xiao, K. Moniz, N. Ghaeli, M. Matus, J. Thompson, E.J. Alm, Quantitative SARS-CoV-2 Alpha Variant B.1.1.7 Tracking in Wastewater by Allele-Specific RT-qPCR, Environ. Sci. Technol. Lett. 8 (2021) 675–682. doi:10.1021/ACS.ESTLETT.1C00375/SUPPL_FILE/EZ1C00375_SI_001.PDF.

[19] Aristotle University of Thessaloniki Research Committee, The Aristotle University of Thessaloniki writes history in dealing with COVID-19 disease, 2021. https://www.rc.auth.gr/Documents/Uploaded/d1792cfe-60a3-4bdf-aeb9-d38c88c8e552.pdf.

[20] M. Petala, D. Dafou, M. Kostoglou, T. Karapantsios, E. Kanata, A. Chatziefstathiou, F. Sakaveli, K. Kotoulas, M. Arsenakis, E. Roilides, T. Sklaviadis, S. Metallidis, A. Papa, E. Stylianidis, A. Papadopoulos, N. Papaioannou, A physicochemical model for rationalizing SARS-CoV-2 concentration in sewage. Case study: The city of Thessaloniki in Greece, Sci. Total Environ. 755 (2021) 142855. doi:10.1016/J.SCITOTENV.2020.142855.

[21] M. Kostoglou, M. Petala, T. Karapantsios, C. Dovas, E. Roilides, S. Metallidis, A. Papa, E. Stylianidis, A. Papadopoulos, N. Papaioannou, SARS-CoV-2 adsorption on suspended solids along a sewerage network: mathematical model formulation, sensitivity analysis, and parametric study, Environ. Sci. Pollut. Res. Int. (2021). doi:10.1007/S11356-021-16528-0.

[22] National Public Health Organization-Greece (EODY), Daily report of epidemiological surveillance of the infection by the new coronavirus (COVID-19), data up to 30 December 2021, 15:00, 2021. https://eody.gov.gr/wp-content/uploads/2021/12/covid-gr-daily-report-20211230.pdf.

[23] National Public Health Organization-Greece (EODY), Daily report of epidemiological surveillance of the infection by the new coronavirus (COVID-19), data up to 31 December 2021, 15:00, 2021. https://eody.gov.gr/wp-content/uploads/2021/12/covid-gr-daily-report-20211231.pdf.

[24] E.I. Agianniotaki, S.C. Chaintoutis, A. Haegeman, K. De Clercq, E. Chondrokouki, C.I. Dovas, A TaqMan probe-based multiplex real-time PCR method for the specific detection of wild type lumpy skin disease virus with beta-actin as internal amplification control, Mol. Cell. Probes. 60 (2021). doi:10.1016/J.MCP.2021.101778.

[25] N.R. Markham, M. Zuker, DINAMelt web server for nucleic acid melting prediction, Nucleic Acids Res. 33 (2005) W577–W581. doi:10.1093/nar/gki591.

[26] Y. You, B.G. Moreira, M.A. Behlke, R. Owczarzy, Design of LNA probes that improve mismatch discrimination, Nucleic Acids Res. 34 (2006). doi:10.1093/nar/gkl175.

[27] R. Owczarzy, Y. You, C.L. Groth, A. V. Tataurov, Stability and mismatch discrimination of locked nucleic acid-DNA duplexes, Biochemistry. 50 (2011) 9352–9367. doi:10.1021/bi200904e.

[28] J.R.C. Pulliam, C. van Schalkwyk, N. Govender, A. von Gottberg, C. Cohen, M.J. Groome, J. Dushoff, K. Mlisana, H. Moultrie, Increased risk of SARS-CoV-2 reinfection associated with emergence of the Omicron variant in South Africa, MedRxiv. (2021) 2021.11.11.21266068. doi:10.1101/2021.11.11.21266068.

[29] WHO, Classification of Omicron (B.1.1.529): SARS-CoV-2 Variant of Concern, (2021). https://www.who.int/news/item/26-11-2021-classification-of-omicron-(b.1.1.529)-sars-cov-2-variant-of-concern (accessed December 27, 2021).

[30] E. Petersen, F. Ntoumi, D.S. Hui, A. Abubakar, L.D. Kramer, C. Obiero, P.A. Tambyah, L. Blumberg, R. Yapi, S. Al-Abri, T. de C.A. Pinto, D. Yeboah-Manu, N. Haider, D. Asogun, T.P. Velavan, N. Kapata, M. Bates, R. Ansumana, C. Montaldo, L. Mucheleng’anga, J. Tembo, P. Mwaba, C.M. Himwaze, M.M.A. Hamid, S. Mfinanga, L. Mboera, T. Raj, E. Aklillu, F. Veas, S. Edwards, P. Kaleebu, T.D. McHugh, J. Chakaya, T. Nyirenda, M. Bockarie, P.S. Nyasulu, C. Wejse, J.-J. Muyembe-Tamfum, E.I. Azhar, M. Maeurer, J.B. Nachega, R. Kock, G. Ippolito, A. Zumla, Emergence of new SARS-CoV-2 Variant of Concern Omicron (B.1.1.529) - highlights Africa’s research capabilities, but exposes major knowledge gaps, inequities of vaccine distribution, inadequacies in global COVID-19 response and control efforts, Int. J. Infect. Dis. 114 (2022) 268. doi:10.1016/J.IJID.2021.11.040.

[31] S.S.A. Karim, Q.A. Karim, Omicron SARS-CoV-2 variant: a new chapter in the COVID-19 pandemic, Lancet (London, England). 398 (2021) 2126. doi:10.1016/S0140-6736(21)02758-6.

[32] S. Wurtzer, P. Waldman, M. Levert, N. Cluzel, J.L. Almayrac, C. Charpentier, S. Masnada, M. Gillon-Ritz, J.M. Mouchel, Y. Maday, M. Boni, V. Marechal, L. Moulin, M. Boni, S. Wurtzer, L. Moulin, J.M. Mouchel, Y. Maday, V. Marechal, S. Le Guyader, I. Bertrand, C. Gantzer, D. Descamps, C. Charpentier, N. Houhou, B. Visseaux, V. Calvez, A.-G. Marcelin, S. Marot, A. Jary, M.-L. Chaix, C. Delaugerre, M. Salmona, J. Legoff, E. Gault, M.-A. Rameix-Welti, A.-S. L’honneur, A.-A. Mariaggi, F. Rozenberg, A.-M. Roque, L. Mouna, H. Fenaux, M. Leruez-Ville, L. Morand-Joubert, S. Lambert-Niclot, A. Schnuriger, C. Dubois, N. Bourgeois-Nicolaos, S. Brichler, H. Delagrèverie, M. Bouvier-Alias, S. Fourati, C. Rodriguez, J.-M. Pawlotsky, SARS-CoV-2 genome quantification in wastewaters at regional and city scale allows precise monitoring of the whole outbreaks dynamics and variants spreading in the population, Sci. Total Environ. 810 (2022) 152213. doi:10.1016/J.SCITOTENV.2021.152213.

[33] ECDC, Threat Assessment Brief: Implications of the emergence and spread of the SARS-CoV-2 B.1.1. 529 variant of concern (Omicron) for the EU/EEA, (2021). https://www.ecdc.europa.eu/en/publications-data/threat-assessment-brief-emergence-sars-cov-2-variant-b.1.1.529 (accessed December 27, 2021).

[34] Public Health England, Investigation of novel SARS-COV-2 variant Variant of Concern 202012/01, 2021. https://assets.publishing.service.gov.uk/government/uploads/system/uploads/attachment_data/file/959438/Technical_Briefing_VOC_SH_NJL2_SH2.pdf.

[35] UK Health Security Agency, Omicron daily overview: 24 December 2021, 2021. https://assets.publishing.service.gov.uk/government/uploads/system/uploads/attachment_data/file/1043866/20211224_OS_Daily_Omicron_Overview.pdf.

[36] Imperial College COVID-19 Response Team, Report 49 - Growth, population distribution and immune escape of Omicron in England, 2021. https://www.imperial.ac.uk/mrc-global-infectious-disease-analysis/covid-19/report-49-omicron/ (accessed December 27, 2021).

[37] UK Health Security Agency, SGTF Epidemic curve data, (2021). https://assets.publishing.service.gov.uk/government/uploads/system/uploads/attachment_data/file/1044330/sgtf_totalepicurve_2021-12-29.csv/preview.

[38] L. Jansen, B. Tegomoh, K. Lange, K. Showalter, J. Figliomeni, B. Abdalhamid, P.C. Iwen, J. Fauver, B. Buss, M. Donahue, Investigation of a SARS-CoV-2 B.1.1.529 (Omicron) Variant Cluster — Nebraska, November–December 2021, MMWR. Morb. Mortal. Wkly. Rep. 70 (2021) 1782–1784. doi:10.15585/MMWR.MM705152E3.

[39] C. del Rio, S.B. Omer, P.N. Malani, Winter of Omicron—The Evolving COVID-19 Pandemic, JAMA. (2021). doi:10.1001/JAMA.2021.24315.

